# Long-term N-Acetylcysteine Treatment Accelerates Improvement in Patient-Reported Outcomes for COVID-19 Patients: A Randomized, Double-Blind, Placebo-Controlled Trial

**DOI:** 10.64898/2025.12.10.25342003

**Authors:** Lifang Li, Lu Ye, Hangzhi He, Chunwei Chai, Yigang Tong, Yanbo Zhang, Hui Zhao

## Abstract

**Objectives:** The coronavirus disease 2019 (COVID-19) pandemic has necessitated the urgent investigation of effective long-term therapeutic agents. This study aimed to evaluate the definitive, long-term causal effect of N-acetylcysteine (NAC) treatment on patient-reported outcomes (PRO) in patients with COVID-19 from hospital admission to six months after discharge.

**Design:** Prospective, multi-center, randomized, double-blind, placebo-controlled trial (RCT).

**Setting:** Five COVID-19-designated hospitals, including the Fourth People’s Hospital, Taiyuan City, China.

**Participants:** Sixty-three confirmed COVID-19 patients were randomly assigned (1:1 ratio) to the NAC-treatment group (n=32) or the placebo control group (n=31). Both groups received conventional treatment. The NAC group received 600 mg oral NAC twice/day, while the control group received an identical-looking placebo. Both regimens were continued post-discharge for a total of six months.

**Main Outcome Measures:** Patients were followed up at discharge and at 1, 3, and 6 months after discharge. Patient-reported outcome was evaluated using the St. George’s Respiratory Questionnaire (SGRQ).

**Results:** Baseline characteristics were balanced between the two groups. Multivariate analysis using the Generalized Estimation Equation (GEE) indicated that NAC treatment, disease severity, and follow-up duration significantly influenced the total SGRQ score. Crucially, the interaction between NAC treatment and follow-up duration was statistically significant (β=-0.436, *P*=0.004).

**Conclusion:** NAC treatment leads to a significantly more rapid decrease in the total SGRQ patient score compared to placebo. This RCT provides causal evidence that the long-term use of NAC accelerates the improvement in the health-related quality of life (HRQoL) of patients recovering from COVID-19.

**Trial Registration:** Evaluation of clinical effect of NAC on lung protection in COVID-19 patients. (www.chictr.org.cn; number: ChiCTR2100049355).

## Introduction

Coronavirus disease 2019 (COVID-19), caused by severe acute respiratory syndrome coronavirus 2 (SARS-CoV-2), has culminated in a global pandemic^1–2^. As of June 4, 2021, over 172 million cases and 3.7 million deaths were confirmed worldwide. Although most patients experience mild-to-moderate symptoms, a large number continue to die, primarily due to acute respiratory distress syndrome (ARDS)^3–4^. Studies suggest that excessive immune activation and cytokine storm are central causes of COVID-19-related lung injury^5–7^. It is currently believed that prolonged oxidative stress, increased production of reactive oxygen species (ROS), and decreased glutathione levels lead to an imbalance in redox homeostasis, which in turn drives excessive immune activation and cytokine storms.

N-acetylcysteine (NAC) is an N-acetyl derivative of the natural amino acid L-cysteine^8^. NAC is widely used clinically, acting as a direct scavenger of ROS to regulate the redox state, modulate the inflammatory response, and exhibit indirect antioxidant properties^9^.

In addition to its antioxidant effects, NAC possesses mucolytic properties. The first COVID-19 death autopsy in China revealed white foamy and jelly-like mucus in the airways, suggesting the presence of highly viscous, difficult-to-discharge secretions that may contribute to patient death^10^. Therefore, the use of expectorant drugs is an important part of adjuvant treatment. The active free sulfhydryl (SH) group in the NAC molecule promotes the breakage of the acid glycoprotein polypeptide disulfide bond (-SS) in sputum^11^, directly splitting DNA and mucin in the hydrolyzed sputum, thereby decomposing respiratory mucus. Clinical studies have shown that NAC treatment improves expectoration difficulty and cough severity^12^.

Furthermore, the protective effect of NAC in influenza and other respiratory viral diseases has been confirmed, showing reduction in the incidence and severity of influenza-like diseases. The mechanism includes inhibiting viral matrix protein expression and regulating the release of pro-inflammatory mediators such as IL-8, IL-6, and TNF-α. Based on its combined anti-oxidative, mucolytic, and anti-inflammatory/antiviral properties, we hypothesize that NAC can play an important role in the treatment of COVID-19.

The St. George’s Respiratory Questionnaire (SGRQ) is a validated, patient-reported instrument used to measure the impact of respiratory diseases and their treatment on a patient’s health-related quality of life (HRQoL)^13^. It includes 50 items divided into three components: symptoms, activities, and impact. The SGRQ total score ranges from 0 to 100 points, with a lower score indicating superior HRQoL. The United States Food and Drug Administration defines a patient-reported outcome (PRO) as any report regarding the status of a patient’s health condition that comes directly from the patient, without interpretation by a clinician or anyone else^14–16^. PRO is a vital measure that reflects the impact of the patient’s disease on their life, independent of survival and physiological indicators.

To our knowledge, previous RCTs have yielded mixed results, primarily focusing on short-term high-dose intravenous NAC in severe patients. To overcome the limitations of previous observational studies and to definitively assess the long-term therapeutic benefit, we conducted a prospective, multi-center, randomized, double-blind, placebo-controlled trial. The primary objective was to evaluate the causal effect of long-term oral NAC use on patient-reported outcomes (PRO) up to six months post-discharge, utilizing the SGRQ as the primary outcome measure.

## Materials and Methods

### Study Design, Randomization, and Blinding

This study was a prospective, multi-center, randomized, double-blind, placebo-controlled trial (RCT), designed and reported in accordance with the CONSORT guidelines.

1. **Randomization:** Eligible patients were randomly assigned (1:1 ratio) to either the NAC-treatment group or the placebo control group using a computer-generated random sequence. The random allocation sequence was generated and concealed by an independent statistician not involved in patient recruitment or outcome assessment.
2. **Blinding:** The study maintained a double-blind design. Neither the participants, the treating physicians, the hospital staff, nor the outcome assessors (those collecting follow-up SGRQ data) were aware of the group assignment. The allocation sequence was sealed in opaque, sequentially numbered envelopes, accessed only in the case of a medical emergency.

### Interventions

Both groups received standard conventional treatment as per national guidelines.

- NAC Treatment Group: Received standard treatment plus oral NAC 600 mg/time, twice daily.
- Placebo Control Group: Received standard treatment plus an identical-looking, identical-tasting placebo at the same dosage and frequency.

Patients in both the NAC and placebo groups continued their assigned oral regimen after discharge until the 6-month follow-up time point.

### Participants

This study was conducted at five designated COVID-19 hospitals, including the Fourth People’s Hospital of Taiyuan City, Shanxi Province. Patients admitted between February and March 2020 were screened for eligibility. The sample size was determined by the number of COVID-19 cases in the region during the study period.

The inclusion criteria were:

a. age > 18 years;
b. SARS-CoV-2-positive real-time reverse transcription polymerase chain reaction tests for nasal swabs or lower respiratory tract samples;
c. chest computed tomography (CT) images confirming pneumonia; and
d. willingness to participate in the study.

The exclusion criteria were:

a. pregnancy and childhood; and
b. inability to provide informed consent.

### Ethics Approval and Informed Consent

The study was approved by the Institutional Review Committee of the Second Hospital of Shanxi Medical University (approval number: [2020] YX number [017]) and the institutional review committees of participating hospitals. Informed consent was obtained during hospitalization.

### Quality Control

Due to the nature of the disease and potential poor compliance with regular follow-up visits, free medication was provided during the follow-up period to enhance patient compliance and minimize bias caused by loss to follow-up. This measure significantly improved patient compliance, with the questionnaire response rate reaching 100%. All staff conducting follow-up visits were uniformly trained.

## Statistical Methods

1. Statistical Description and Correlation: Measurement data are presented as mean values ± standard deviations (X ̅±S). Composition ratio was used to describe counted data. Pearson’s correlation coefficient was used to evaluate and test the correlations between symptoms, activity, impacts, and total scores on the SGRQ scale.
2. Univariate Analysis: The Mann–Whitney *U* test was used to compare the differences in SGRQ component and total score changes between the NAC-treatment and non-NAC-treatment groups at each follow-up time point (compared with the score at hospital discharge). The Kruskal–Wallis *H* test was used to analyze the changes in SGRQ scores at all follow-up time points within each group.
3. Multivariate Analysis: The Generalized Estimation Equation (GEE) was used to perform a multi-factor analysis^17^ to minimize the influence of confounding factors on the SGRQ score. To improve the clinical interpretation of model parameters and reduce variable dispersion, the unit value of difference (UVD) of the corresponding variable was constructed based on the Minimal Clinically Important Difference (MCID)^27^. For the total score, its UVD is:

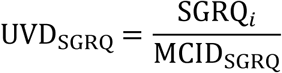

In the above formula, the SGRQi is the total SGRQ score at follow-up time *i*, and the MCID_SGRQ_ is the MCID of the total score. According to previous research reports by scholars, the MCID of the total score is usually set to 4 points. The UVD_SGRQ_ was incorporated as a dependent variable into the generalized estimation equation for analysis. Its meaning in the generalized estimation model was as follows: when other variables remain unchanged, the total score increases (or decreases) by several units due to changes in certain factors. Further, when the absolute value of the UVD_SGRQ_ change in the model exceeds 1, it often suggests that the change in the total score may have practical clinical significance.

4. Data Analysis Software

EpiData software (version 3.1) was used for data double-entry and verification. All statistical analyses were performed using SPSS (version 25.0; IBM, Armonk, NY, USA), with the inspection level set at α = 0.05.

## Results

### 1. Baseline Status and Recruitment

A total of 63 confirmed COVID-19 patients were enrolled and randomized into the study. The mean age of participants was 44.65±19.46 years, and the average length of hospital stay was 16.24±7.79 days. Males constituted 60.3% of the cohort, and 71.43% were diagnosed with a light illness upon admission.

Due to the successful implementation of the randomization procedure, the NAC-treatment group (n=32) and the placebo control group (n=31) were well-balanced and comparable across all baseline demographic and clinical characteristics, including age, gender, BMI, comorbidities, and initial disease severity (Table 1). No statistically significant differences were found between the two groups at baseline (*P*>0.05,for all comparisons).

**Table 1.**
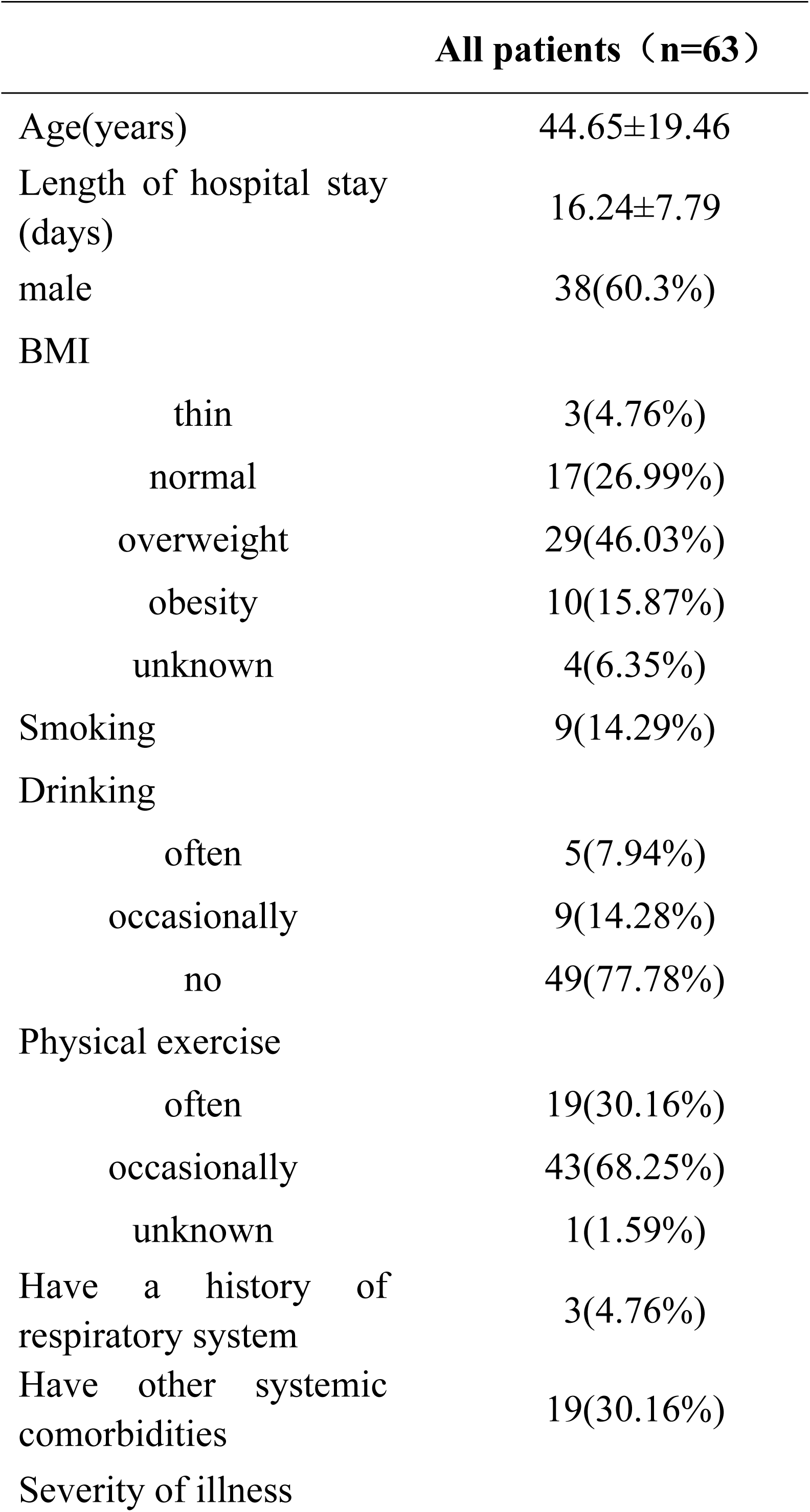
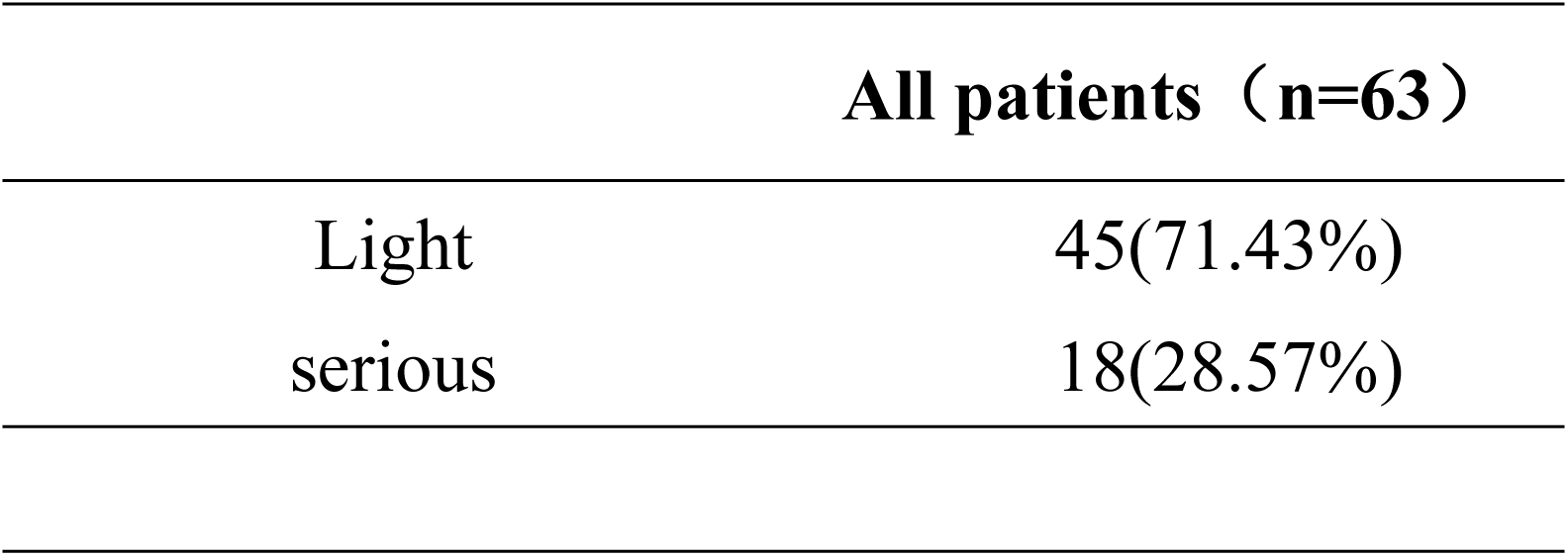
Baseline situation of patients with COVID-19.

### 2. Correlation Analysis of SGRQ Scores

The Pearson correlation coefficient analysis was used to evaluate the correlation between SGRQ component scores. Strong correlations were found between the scores of the three SGRQ components (Symptoms, Activity, and Impacts) and between each component score and the total score (*P* < 0.001). The detailed correlation matrix is shown in Table 2.

**Table 2.**
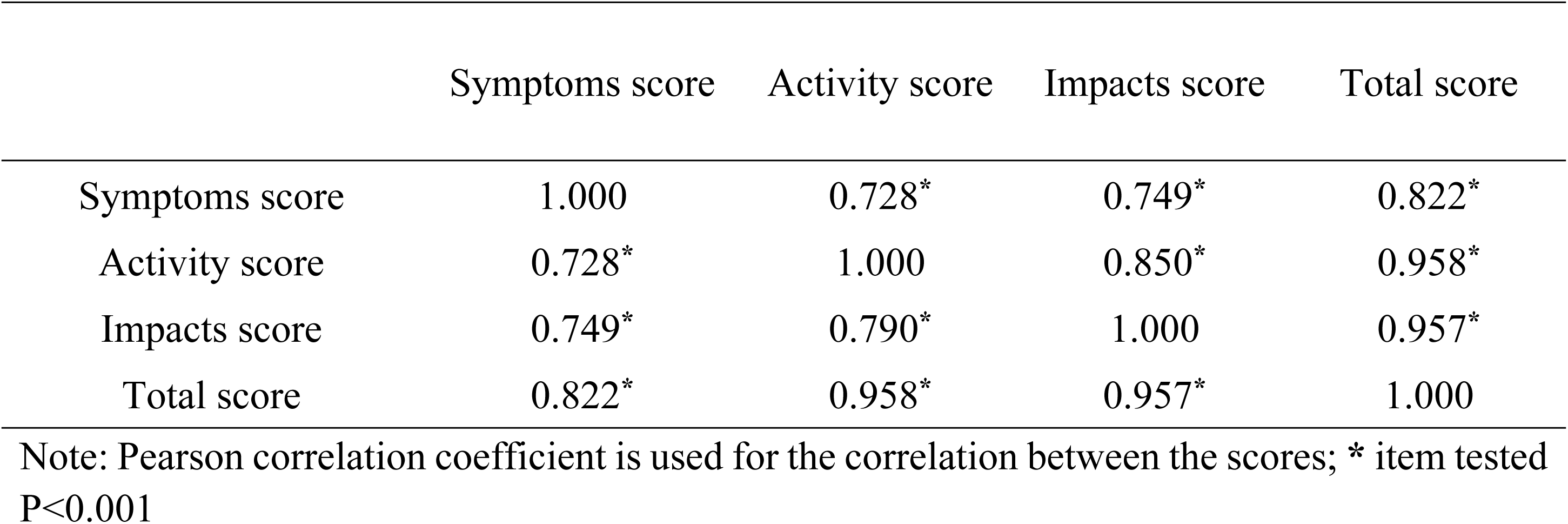
Correlations of SGRQ series scores in all patients.

|  | Symptoms score | Activity score | Impacts score | Total score |
| --- | --- | --- | --- | --- |
| Symptoms score | 1.000 | 0.728* | 0.749* | 0.822* |
| Activity score | 0.728* | 1.000 | 0.850* | 0.958* |
| Impacts score | 0.749* | 0.790* | 1.000 | 0.957* |
| Total score | 0.822* | 0.958* | 0.957* | 1.000 |

Note: Pearson correlation coefficient is used for the correlation between the scores; \* item tested P<0.001

Specifically, the total score exhibited the strongest correlation with the Activity score (r=0.958), followed closely by the Impacts score (r=0.957) and the Symptoms score (r=0.822).

### 3. Univariate Analysis: Group Differences in SGRQ Change Scores

Univariate analysis (Mann–Whitney U test) compared the median change differences in SGRQ scores (relative to discharge) between the NAC-treatment group and the placebo control group at each follow-up time point (Table 3). The results showed statistically significant greater improvements in the NAC-treatment group compared to the placebo control group:

- 1 Month Post-Discharge: Significant differences were found in the Impacts score (*P*<0.001) and Total score (*P*=0.042).
- 3 Months Post-Discharge: Significant differences were observed in the Activity score (*P*=0.022), Impacts score (*P*<0.001), and Total score (*P*=0.004).
- 6 Months Post-Discharge: The significant differences persisted across the Activity score (*P*=0.022), Impacts score (*P*<0.001), and Total score (*P*=0.002).

**Table 3.** Changes in SGRQ scores of the two groups of COVID-19 patients at each follow-up time.

|  | **NAC**<br>**treatment**<br>**(n=32)** | **Without NAC**<br>**treatment**<br>**(n=31)** | *U* | *P* |
| --- | --- | --- | --- | --- |
| **Change of one month after discharge compared with discharge** |  |  |  |  |
| Symptoms score | 13.10±17.32 | 7.60±12.94 | 555.000 | 0.411 |
| Activity score | 12.12±15.72 | 8.74±11.95 | 573.500 | 0.281 |
| Impacts score | 12.12±17.80 | 2.24±9.44 | 753.000 | **<0.001** |
| Total score | 12.28±15.54 | 5.08±10.05 | 644.000 | **0.042** |
| **Change of three month after discharge compared with discharge** |  |  |  |  |
| Symptoms score | 18.06±18.25 | 10.04±14.94 | 632.000 | 0.061 |
| Activity score | 24.19±27.52 | 11.70±15.21 | 660.500 | **0.022** |
| Impacts score | 15.39±21.01 | 2.41±10.04 | 800.000 | **<0.001** |
| Total score | 18.49±21.25 | 6.80±11.99 | 707.000 | **0.004** |
| **Change of six month after discharge compared with discharge** |  |  |  |  |
| Symptoms score | 18.53±19.66 | 11.22±13.97 | 612.500 | 0.108 |
| Activity score | 24.19±27.52 | 11.70±15.21 | 660.500 | **0.022** |
| Impacts score | 15.57±21.02 | 2.74±10.29 | 793.000 | **<0.001** |
| Total score | 18.65±21.38 | 6.33±12.23 | 723.000 | **0.002** |

Note: Mann-Whitney *U* test was used for the comparison of differences in score changes between groups

Within-group longitudinal comparison using the Kruskal–Wallis H test demonstrated significant improvements across all follow-up time points in both groups (Table 4), indicating continuous recovery after discharge. Notably, the NAC-treated group showed a more pronounced and accelerated reduction in SGRQ total, activity, and impacts scores over time.

**Table 4.**
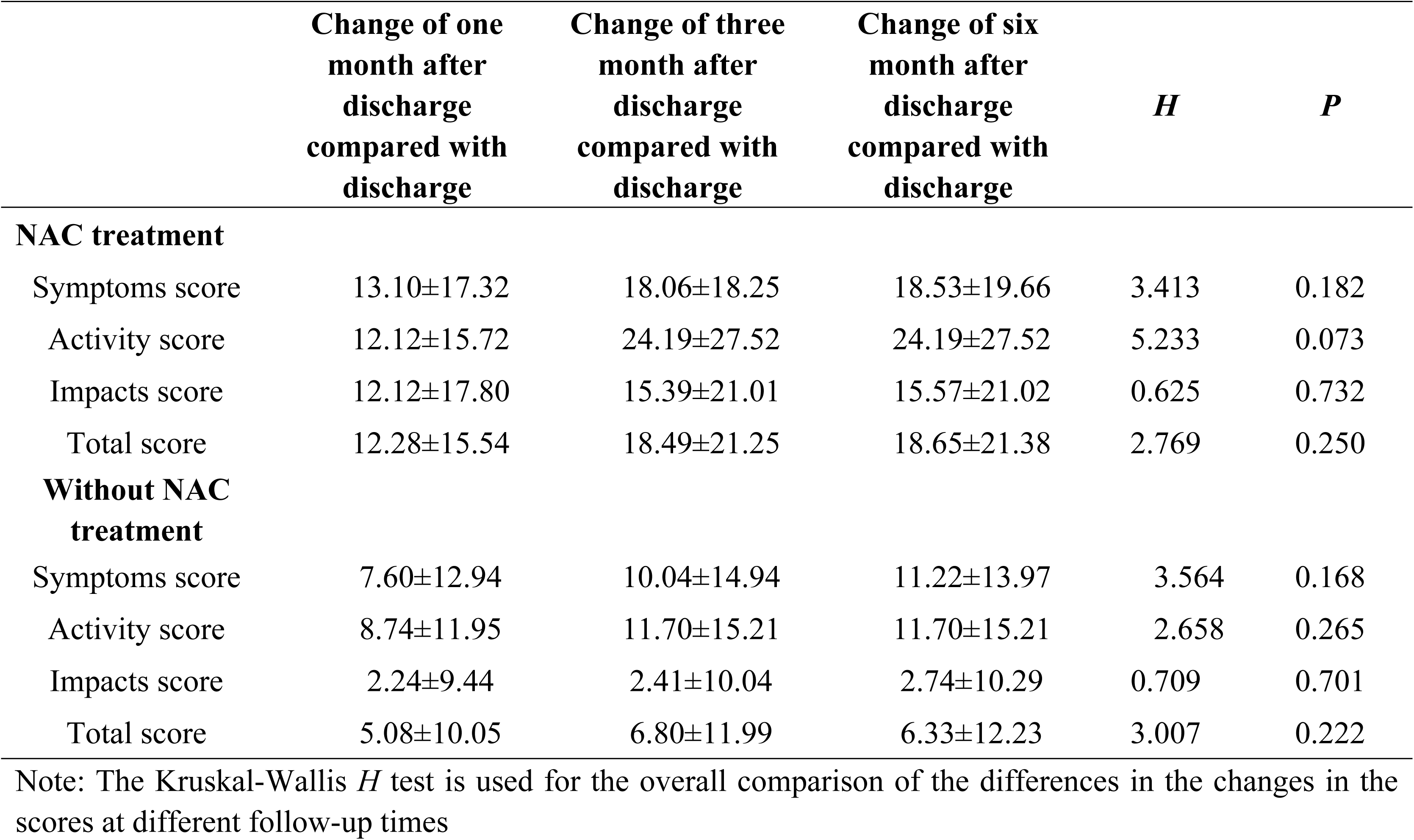
Changes in SGRQ scores of patients at each follow-up time in the two groups.

### 4. Multivariate Analysis: Factors Influencing SGRQ Improvement Rate

Multivariate analysis utilizing the Generalized Estimation Equation (GEE) model was performed to analyze the factors influencing the change in the SGRQ total score UVD_SGRQ_ over time. GEE analysis showed that NAC treatment, disease severity, and follow-up duration were all statistically significant predictors (Figure 2).

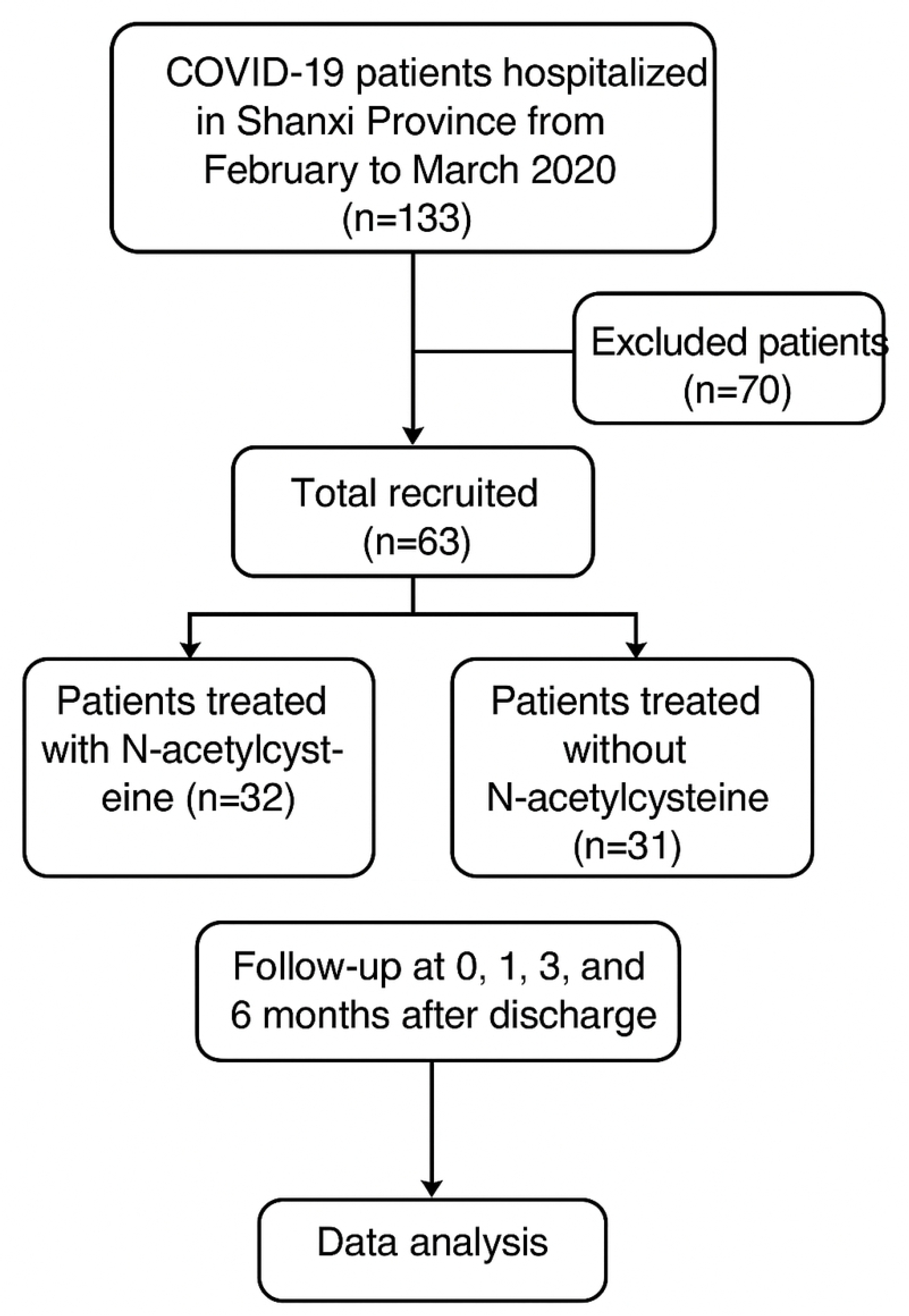

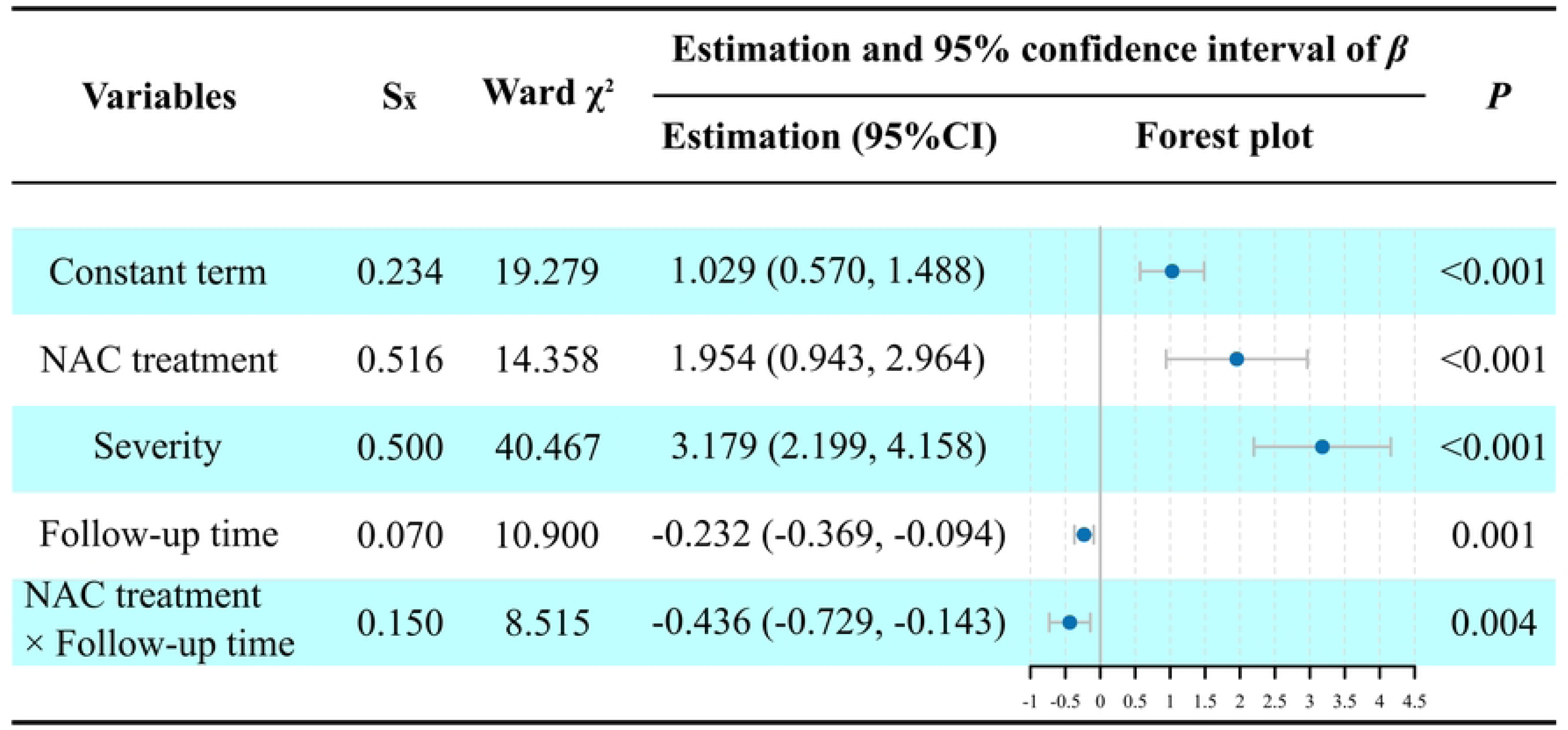

Crucially, the interaction between NAC treatment and follow-up duration was statistically significant (β=-0.436, *P*=0.004) (Figure 2). This negative β value for the interaction term provides causal evidence that NAC treatment significantly accelerated the rate of SGRQ total score decrease compared to placebo.

### 5. Visual Trend Confirmation

The visual trend of the marginal mean SGRQ total score confirmed the analytical findings (Figure 3). The slope of the average change in the total score for the NAC-treated group was significantly steeper than that for the placebo control group over the six-month period. This demonstrates that long-term NAC treatment accelerated the improvement in the health-related quality of life of COVID-19 convalescents.

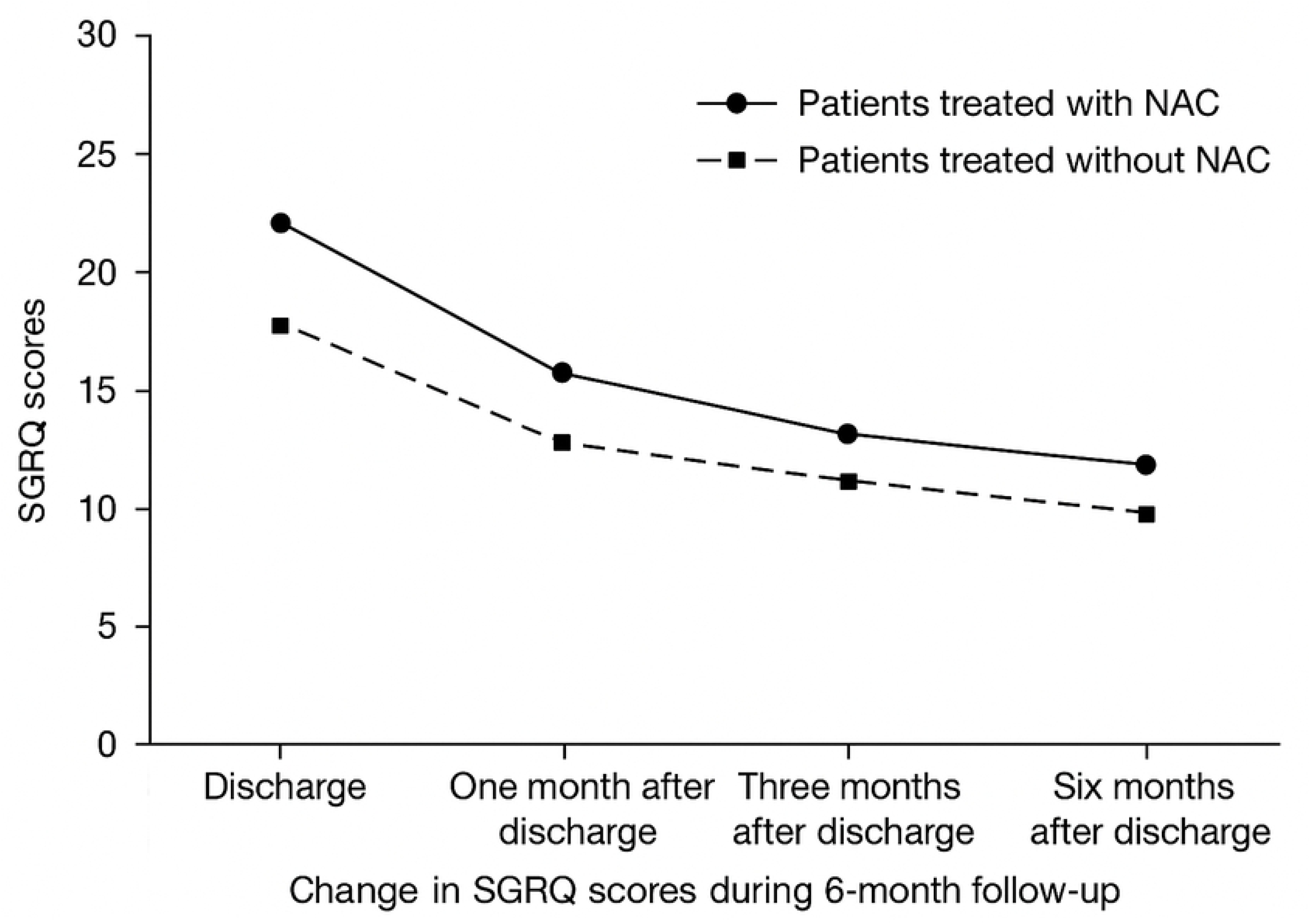

## Discussion

NAC is widely utilized in the management of respiratory diseases, owing to its established anti-oxidation, mucolytic, and potential virus-inhibiting properties^18–20^. Numerous scientific reviews have demonstrated the biochemical and physiological rationale for its use as an adjuvant therapy for COVID-19. Furthermore, supportive evidence comes from case reports, such as its role in improving refractory hypercapnia through airway management in critical COVID-19 patients and improving clinical symptoms in G6PD-deficient patients with severe COVID-19^21^.

However, clinical evidence from initial randomized controlled trials (RCTs) has been mixed. Previous trials have often focused on high-dose, short-term intravenous administration. For instance, a meta-analysis showed that NAC may reduce mortality^22^. Conversely, other trials/meta-analyses have found that high-dose NAC did not significantly affect disease progression ^23,24^. Similarly, a pilot study on intravenous NAC in patients with mild-to-moderate COVID-19-related ARDS showed improvement in clinical status but failed to achieve statistical significance^25^, a finding echoed by another RCT^26^.

Our study addresses this gap by employing a robust randomized, double-blind, placebo-controlled design and focusing on the long-term impact of oral NAC on patient-reported outcomes (PRO) over a six-month recovery period. Our univariate analysis showed that, compared to placebo, the NAC-treated group achieved statistically significant and clinically meaningful improvements across the SGRQ components (Activity, Impacts, and Total scores) at 3 and 6 months post-discharge (Table 3).

The most compelling finding, however, stems from the multivariate Generalized Estimation Equation (GEE) analysis: the statistically significant interaction term between NAC treatment and follow-up duration (β=-0.436, *P*=0.004). This interaction term provides strong causal evidence that the rate of improvement (decrease in SGRQ total score) in the NAC-treated group was significantly faster than in the placebo control group. Figure 3 visually confirms this accelerated trajectory, showing a steeper slope for the NAC group over time. This suggests that while NAC may not be effective in rapidly reversing acute, severe pathology, its sustained use is highly effective in driving long-term functional and quality-of-life recovery, which is a significant patient-centric outcome.

The accelerated recovery observed in the NAC group is biologically plausible in the context of post-COVID-19 recovery. The persistence of symptoms often categorized as "long COVID" is widely attributed to protracted systemic inflammation and chronic oxidative stress. By sustaining the supply of cysteine and promoting the replenishment of glutathione (GSH)—the body’s master antioxidant—the long-term oral NAC regimen likely maintained a more favorable redox environment. This sustained mechanism appears critical for mitigating the chronic inflammatory damage and facilitating the repair and functional restoration of the respiratory system and overall well-being, surpassing the body’s natural recovery rate seen in the placebo group.

## Limitations

Despite the robust RCT design, this study has certain limitations. First, the sample size remains relatively modest, which, while sufficient for detecting the primary outcome based on the GEE interaction term, limits the power for subgroup analysis. Second, all samples were limited to one province due to objective restrictions. Hence, the research results may exhibit regional characteristics. Future research should expand the sample sources to multiple provinces and increase the sample size to further improve the representativeness and reliability of the research findings.

## Conclusion

NAC treatment significantly accelerates the rate of decrease in the total SGRQ patient score compared to placebo. The findings from this randomized controlled trial suggest that the long-term use of NAC provides a substantial and demonstrable benefit by accelerating the improvement of health-related quality of life in patients recovering from COVID-19.

## Data Availability

Data cannot be shared publicly because of patient confidentiality and ethical restrictions imposed by the Institutional Review Board (IRB) of the Second Affiliated Hospital of Shanxi Medical University. Data are available from the Second Affiliated Hospital of Shanxi Medical University Research Data Access Committee (contact via for Data Access Inquiries) for researchers who meet the criteria for access to confidential patient data. The data for this study are held by Research Manager.

http://www.medresman.org.cn

## Acknowledgements

We acknowledge all the patients who participated in this trial. We are grateful for the support provided by the Emergency Scientific Research Project for the Prevention and Control of the Novel Coronavirus Outbreak in Shanxi Province (Project No: 202003D32003/GZ).

## Conflict Of Intersts

None declared.

## Ethical Approval

The study was approved by the Institutional Review Committee of the chief investigator’s hospital (the Second Hospital of Shanxi Medical University, China (No.2020YX017) and the institutional review committees of participating hospitals. According to the approval of the Local Ethics Committee, informed consent was obtained during hospitalization.

## Author Contribution

All authors have made a substantial contribution to the research design and approved the final version of this paper. Hui Zhao design the work, Lifang Li drafted the paper, Yanbo Zhang and Yigang Tong revised the article critically, Hangzhi He analyzed the data, Chunwei Chai and Lu Ye acquired the data. All authors have read and approved the final manuscript.

## Data Availability Statement

The data for this study are held by Research Manager (http://www.medresman.org.cn).

## Notes

### Competing Interest Statement

The authors have declared no competing interest.

### Clinical Trial

ChiCTR2100049355

### Clinical Protocols

http://www.chictr.org.cn

### Funding Statement

Yes

### Author Declarations

The study was approved by the Institutional Review Committee of the chief investigator’s hospital (the Second Hospital of Shanxi Medical University, China (No.2020YX017) and the institutional review committees of participating hospitals.

## References

1. Johns Hopkins University Coronavirus Resource Center. COVID-19 dashboard by the Center for Systems Science and Engineering (CSSE) at Johns Hopkins University. 2021. https://coronavirus.jhu.edu/map.html.

2. Polack FP, Thomas SJ, Kitchin N, et al. Safety and Efficacy of the BNT162b2 mRNA Covid-19 Vaccine. N Engl J Med. 2020. 383(27): 2603–2615.

3. Cao B, Wang Y, Wen D, et al. A Trial of Lopinavir-Ritonavir in Adults Hospitalized with Severe Covid-19. N Engl J Med. 2020. 382(19): 1787–1799.

4. Rojas-Marte G, Khalid M, Mukhtar O, et al. Outcomes in patients with severe COVID-19 disease treated with tocilizumab: a case-controlled study. QJM. 2020. 113(8): 546–550.

5. Huang C, Wang Y, Li X, et al. Clinical features of patients infected with 2019 novel coronavirus in Wuhan, China. The Lancet. 2020. 395.

6. Meftahi GH, Jangravi Z, Sahraei H, Bahari Z. The possible pathophysiology mechanism of cytokine storm in elderly adults with COVID-19 infection: the contribution of "inflame-aging". Inflamm Res. 2020. 69(9): 825–839.

7. Soy M, Keser G, Atagündüz P, Tabak F, Atagündüz I, Kayhan S. Cytokine storm in COVID-19: pathogenesis and overview of anti-inflammatory agents used in treatment. Clin Rheumatol. 2020. 39(7): 2085–2094.

8. Mokhtari V, Afsharian P, Shahhoseini M, Kalantar M, Moini A. A Review on Various Uses of N-Acetyl Cysteine. CELL J. 2017. 19: 11.

9. Poe FL, Rangel S. N-Acetylcysteine to Combat COVID-19: An Evidence Review. Ther Clin Risk Management. 2020. 16: 1047–1055.

10. Liu Q WR, Qu G WY, Liu P ZY, Fei G RL and Zhou Y LL. Gross examination report of COVID-19 death autopsy. Journal of Forensic Medicine. 2020. 36: 21–23.

11. Aldini G, Altomare A, Baron G, et al. N-Acetylcysteine as an antioxidant and disulphide breaking agent: the reasons why. Free Radic Res. 2018. 52(7): 751–762.

12. Qi Q, Ailiyaer Y, Liu R, et al. Effect of N-acetylcysteine on exacerbations of bronchiectasis (BENE): a randomized controlled trial. Respir Res. 2019. 20(1): 73.

13. Lo K, Donohue J, Judson M, Wu Y, Barnathan E, Baughman R. The St. George’s Respiratory Questionnaire in Pulmonary Sarcoidosis. LUNG. 2020. 198.

14. Administration USFaD. Guidance for industry: patient-reported outcome measures:Use in medical product development to support labeling claims. 2009-9. http://www.fda.gov/downloads/Drugs/GuidanceComplianceRegulatoryInformation/Guidances/UCM193282.pdf.

15. Jiang Q. Patient-reported outcome and its application in hematological neoplasm. Zhonghua Xue Ye Xue Za Zhi. 2019. 40: 614–619.

16. Afroz N, Gutzwiller FS, Mackay AJ, Naujoks C, Patalano F, Kostikas K. Patient-Reported Outcomes (PROs) in COPD Clinical Trials: Trends and Gaps. Int J Chron Obstruct Pulmon Dis. 2020. 15: 1789–1800.

17. Harrell FE Jr. Regression modelling strategies with application to linear models, logistic regression, and survival analysis. 2001. 1. New York. Springer-Verlag New York.

18. Van Hezik EJ. N-acetylcysteine for prevention and treatment of COVID-19: Current state of evidence and future directions. Future Sci OA. 2025.

19. De Flora S, Balansky R, La Maestra S. Rationale for the use of N-acetylcysteine in both prevention and adjuvant therapy of COVID-19. FASEB J. 2020. 34(10): 13185–13193.

20. Dominari A, Hathaway Iii D, Kapasi A, et al. Bottom-up analysis of emergent properties of N-acetylcysteine as an adjuvant therapy for COVID-19. World J Virol. 2021. 10(2): 34–52.

21. Liu Y, Wang M, Luo G, et al. Experience of N-acetylcysteine airway management in the successful treatment of one case of critical condition with COVID-19: A case report. Medicine (Baltimore). 2020. 99(42): e22577.

22. Xu D, Li Y, Guo X, et al. Effect of N-Acetylcysteine on mortality in COVID-19 patients: A systematic review and meta-analysis of randomized controlled trials. Springer Nature Switzerland AG. 2025.

23. Julio Cesar Garcia de Alencar CdLM, Alicia Dudy Müller CEC, Marina Akemi Fukuhara EAdS, et al. Double-blind, randomized, placebo-controlled trial with N-acetylcysteine for treatment of severe acute respiratory syndrome caused by COVID-19. CLIN INFECT DIS. 2021. 1(72): 11.

24. Wang P, Hou Y, Yu H, et al. Is N-acetylcysteine effective in treating patients with coronavirus disease 2019? A meta-analysis. J Clin Med. 2023. 12(4): 1599.

25. Taher A, Lashgari M, Sedighi L, Rahimi-Bashar F, Poorolajal J, Mehrpooya M. A pilot study on intravenous N-Acetylcysteine treatment in patients with mild-to-moderate COVID19-associated acute respiratory distress syndrome. Pharmacol Rep. 2021 : 1–10.

26. Shayan N, Emadi S, Alijanpour S, et al. Efficacy of N-acetyl Cysteine in Severe COVID-19 Patients: A Randomized Controlled Phase III Clinical Trial. J Janandeesar Navandeesan Pars. 2023. 3(2): e129817.

27. Jones PW. Interpreting thresholds for a clinically significant change in health status in asthma and COPD. Eur Respir J. 2002. 19(3): 398–404.

